# INFLUENCE OF MOLECULAR GENETIC CLASSES ON BEHAVIOR IN PRADER-WILLI SYNDROME

**DOI:** 10.1101/2025.11.23.25340826

**Authors:** Ranim Mahmoud, Merlin G. Butler, Wai Park, Jennifer L. Miller, Daniel J. Driscoll, June-Anne. Gold, Virginia. Kimonis

## Abstract

A wide range of behavioral phenotypes has been described in PWS patients including autism spectrum disorder (ASD). The prevalence of behavioral disorders was studied in 292 participants over 3 years with genetically confirmed PWS (N=164 females and N=128 males) with deletion (N=182) and UPD (N=99). The prevalence of ASD, and other behavioral disorders was tested for association with gender, genetic subtypes, and growth hormone (GH) treatment. The prevalence of ASD in PWS individuals was 19.5%, in concordance with previous studies at 25%. The frequency of ADHD was 10.7%. The mean age at diagnosis for ASD, ADHD, and disruptive behavior was 14.9±10.5, 9.3±5.9, and 19.6±14 years, respectively. There was no statistically significant difference in the prevalence of ASD and ADHD between deletion and UPD subjects, and between GH-treated and non-treated subjects. Patients with deletions had higher frequencies of anxiety than those with UPD(p=0.006). GH-treated participants had a lower frequency of depression and a higher frequency of anxiety than non-treated participants (p=0.04, p=0.02, respectively). This is the largest study to evaluate an association between genetically confirmed PWS and ASD. We found no significant difference in the frequency of ASD and other behavioral disorders across the genetic groups and GH treatment.

## Introduction

Prader-Willi syndrome (PWS) is a genetic neurodevelopmental disorder caused by the absence of paternally expressed imprinted genes. PWS has three genetic classes deletion in about 70% of cases due to deletion of the proximal long arm of chromosome 15 (15q11.2-q13). Maternal uniparental disomy (mUPD), which occurs in 27% of cases in which the patient has two copies of chromosome 15s are inherited from the mother and imprinting defect in about 3% of cases. PWS is characterized by hypotonia in the neonatal period, difficulty of feeding during infancy, followed by hyperphagia, and morbid obesity in childhood. Short stature, intellectual disability (average IQ: 60–70), delayed motor and language development, and behavioral problems aseasy frustration, impulsive, quick anger, stubborn and inflexible. They have a high pain threshold, highly anxious and prone to skin-picking and other obsessive-compulsive behavior (Butler, 2023; Bittel, & Butler, 2005; Burman., et al 2001; Butler, 1990). PWS is one of several genetic disorders associated with autism such as Rett syndrome, Fragile-X syndrome, Smith-Lemli-Opitz syndrome, Tuberous sclerosis, and Smith Magenis syndrome (Butler et al.,2012). Variation in the molecular mechanism of the disorder can result in significant differences in the clinical presentation mainly affecting behavioral and psychiatric phenotype (Butler et al.,2004; Zarcone et al., 2007; Dykens, & Roof, 2008; Yang et al.,2013; Butler et al., 2019). Patients with DEL are more likely to have serious behavioral problems, such as self-injury, food-stealing, and compulsive behaviors. On the other side Individuals with UPD tend to have significantly higher verbal IQ scores and are more prone to develop psychotic disorders (Grugni, & Marzullo,2016).

The prevalence of autism spectrum disorders (ASD) in patients with PWS was estimated to be 25.3%. The PWS 15q11–q13 region contains ASD susceptibility genes. Due to the duplication and overexpression of maternally expressed genes in the 15q11–q13 region patients with Maternal UPD had higher risk for ASD than those with deletion (Wassink, & Piven, 2002). Veltman et al., 2005 reported that 38% of patients with mUPD had associated ASD in comparison to 18% of PWS with deletion. The etiology of ASD is complex and involves genetic factors and environmental factors (epigenetics). Patients with ASD have cognitive impairment, ranging from profound intellectual disability to above-average intellectual functioning and language disability, ranging from absent speech to fluent language. Also, they have impairments in social interaction, communication, and stereotyped repetitive and compulsive behaviours (Dykens et al.,1996; Clarke et al.,2002).

Recombinant growth hormone treatment (GH) for Prader-Willi syndrome was approved in the United States in 2000 and has since been widely recognized as a beneficial treatment for the co-morbidities associated with the syndrome. GH improves linear growth, body composition and lean muscle mass, metabolism and energy expenditure, bone mineral density in PWS patients In addition to the physical improvements, GHT may improve behavior and cognition (Butler et al., 2019; Myers et al.,2007; Kucharska et al.,2024; Höybye et al.,2005; Butler et al.,2022). The purpose of this study was to describe differences in PWS behaviors, by molecular subtype, age groups, sex, and growth hormone treatment.

## Methods

Data from 355 individuals with genetically confirmed PWS were collected at the University of California, Irvine, California; University of Florida Health Science Center, Gainesville, Florida; University of Kansas Medical Center, Kansas City, Kansas; and Vanderbilt University Medical Center, Nashville, Tennessee and entered into the National Institute of Health (NIH) funded Rare Disease Clinical Research Network (RDCRN) PWS registry Written informed consent was obtained from the participants or guardians prior to enrollment using approved human subjects research consent forms at the four sites. Using retrospective guardian-reported data from the NIH Rare Disease Clinical Research Network (RDCRN) natural history of PWS, we compared autism spectrum disorders (ASD), attention deficit hyperactive disorder (ADHD) and disruptive behavior disorder in genetically confirmed PWS patients by molecular cytogenetic testing. This study used the largest dataset on patients with PWS to explore the association between PWS genetic classes and growth hormone treatment with the prevalence of eight psychiatric behaviors (depressed mood, anxiety, skin picking, nail picking, compulsive counting, compulsive ordering, plays with strings, visual hallucinations, and delusions.

The data were summarized using mean and standard deviation (SD) for continuous variables. Subject groups were subdivided by PWS molecular genetic classes and growth hormone treatment. From a cohort of 355 individuals with PWS, 292 participants over three years with DEL or mUPD classes were analyzed. Patients were categorized into three age groups, namely 3-12 years (N= 139), 12-18 years (N= 61), and older than 18 years (N= 92). The statistical analyses were performed using Statistical Package for Social Sciences (SPSS) 29 Statistics software (Armonk, NY, USA). We used a p value of .05 for statistical significant associations using Pearson Chi-Square tests between the behavior phenotypes and genetic classes. The imprinting center group was excluded in the analysis because of the small cohort (5% of the PWS subjects).

## Results

The study included 292 participants, with 164 (56%) females and 128 (44%) males. There were 182 (62%) participants with 15q11.2-q13 deletion and 99 (34%) with UPD. Sixty-two percent were actively on GH treatment (n=181). The overall prevalence of ASD in individuals with PWS in our study was (19.5%), in agreement with previous studies (25%). The frequency of ADHD in this cohort was (12%). The mean age of diagnosis for ASD and ADHD and disruptive behavior disorder was (15.7±10.3, 10± 6.3, 19.6±14) years; respectively. Delusion was present in 8.6%, depression in 16.3%, anxiety in 62%, skin picking in 74%, visual hallucinations in 10%, compulsive counting in 22%, play with string in 33%, compulsive disorders in 53%, and oppositional defiant in 6%.

When comparing deletion with UPD, there was no statistically significant difference in the prevalence of ASD and ADHD between the deletion and UPD subjects (p=0.56 and p=0.65; respectively. The frequencies for psychiatric or behavioral disorders in those with deletion versus UPD were not significantly different for compulsive counting (14.7% vs.8%; p=0.53), compulsive ordering (33% vs. 19%; p=0.43), playing with strings (21% vs.11%, p=0.43), depression (17% vs 15%, p=0.75), visual hallucinations (6% vs. 3%; p=0.33), and delusions (5% vs. 3%; p=0.48). Participants with deletions had higher frequencies of anxiety (34% vs.29%, p= 0.005) and skin picking (49% vs.26%, p=0.009) than those with UPD. When comparing the three age groups we found that Visual hallucinations, depression, and skin picking were significantly higher in the older age group (p=0.016, <0.001, 0.008), respectively. Anxiety was significantly higher in participants from 12-18 years (p=0.006) (Table 1).

**Table 1:** Differences in PWS behavior by age groups.

| Variables | 3-12years (N=139) | 12-18 years (N=61) | >18 years (N=92) | P value |
| --- | --- | --- | --- | --- |
| Visual hallucinations | 7 (5%) | 3 (5%) | 15 (16%) | 0.016 |
| Delusion | 10 (7%) | 3 (5%) | 10 (11%) | 0.508 |
| Depression | 6 (4%) | 15 (24.5%) | 28 (30%) | <0.001 |
| Anxiety | 79 (56%) | 60 (98%) | 45 (49%) | 0.006 |
| Skin picking | 90 (65%) | 47 (77%) | 80 (87%) | 0.008 |
| **Compulsive counting** | 37 (27%) | 13 (21%) | 25 (27%) | 0.301 |
| **Play with strings** | 48 (34.5%) | 24 (39%) | 25 (27%) | 0.428 |
| **Compulsive disorder** | 75 (53%) | 31 (51%) | 43 (47%) | 0.882 |

**Table 2:**
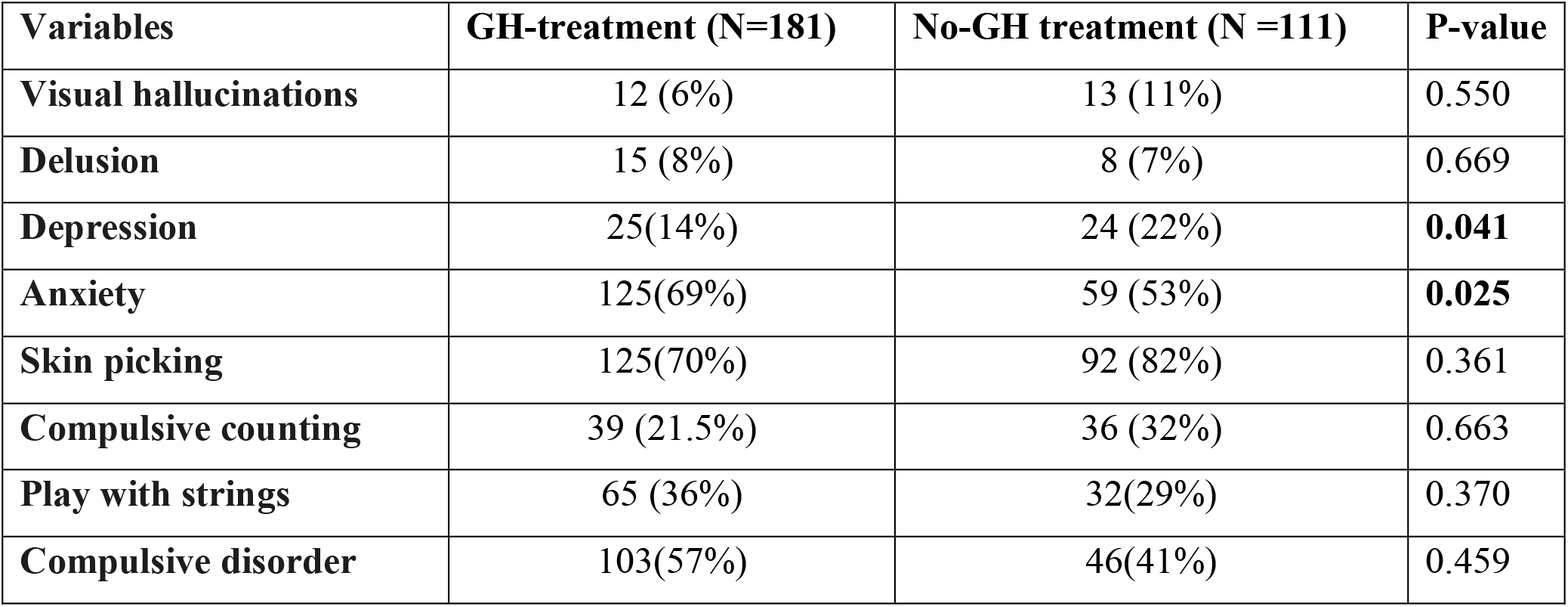
Differences in PWS behavior by GH treatment.

The study participants were analyzed according to their history of growth hormone (GH) treatment (treated vs. not treated) regardless of their PWS molecular class. We found that 62% of patients received GH treatment. The mean age at which GH treatment began was 3.8± 6 years. We found that GH treated patients had a lower frequency of depression and a higher frequency of anxiety than those not treated with GH. No statically significant difference was found in ASD and ADHD between GH treated and non-GH treated.

### Comparison of Effect of Growth Hormone Treatment on Specific PWS Molecular Genetic Classes

Analysis of effects of growth hormone (GH) treatment was also investigated for each individual molecular genetic class (Table 3). The mean age of starting GH treatment was 3.8 ± 6 years (range from birth to 49 years) with an average duration of 13 ± 0.8 years (range from birth to 53 years). Behavioral disorders were studied in the molecular classes; GH treated individuals vs non-GH-treated with the deletion group and UPD group on GH treatment or non-GH treatment. We found that patient with deletion who received GH treatment had lower frequency of skin picking (42.5% vs. 57%; p= 0.002), and depression (9% vs 13.5%; p=0.041) than those without GH treatment. Patients with deletion who received GH treatment had higher frequency of anxiety than those without GH treatment (37%vs. 28%; p=0.032). There was no statistically significant difference in behavioral disorders in GH-treated vs non-GH treated in the UPD group.

**Table 3:** Effect of Growth Hormone Treatment on Specific PWS Molecular Genetic Classes.

| Variables | Deletion |  | P-value | UPD |  | P-value |
| --- | --- | --- | --- | --- | --- | --- |
|  | GH-treated | Non-GH-treated |  | GH-treated | Non-GH-treated |  |
| Visual hallucinations | 9 (5%) | 7 (6%) | 0.792 | 3 (2%) | 6 (5%) | 0.741 |
| Delusion | 11 (6%) | 4 (3%) | 0.571 | 4 (2.5%) | 4 (4%) | 0.470 |
| Depression | 16 (9%) | 15 (13.5%) | **0.041** | 9 (5%) | 9 (8%) | 0.234 |
| Anxiety | 67 (37%) | 32 (28%) | **0.032** | 58 (32%) | 27 (24%) | 0.523 |
| Skin picking | 77 (42.5%) | 63 (57 %) | **0.002** | 48(26.5%) | 29 (26%) | 0.184 |
| Compulsive counting | 25 (14%) | 26 (23%) | 0.883 | 14 (7%) | 11(9%) | 0.342 |
| Play with strings | 35 (19%) | 26 (23%) | 0.893 | 30 (6.5%) | 6 (5%) | 0.064 |
| Compulsive disorder | 59 (32.5%) | 35 (31.5%) | 0.585 | 44 (24%) | 11(9%) | 0.654 |

### Age of start of GH treatment and PWS behavioral disorders

Univariate analysis was done to identify association between age of start of GH with depression and anxiety. PWS patients who had depression started GH treatment at a later age than those without depression with median age of 7 years and a range from 4 months-49 years vs 1.5 years range from birth to 40 years; p=0.003. Early initiation of GH treatment was associated with a lower risk of depression (p=0.003) without increasing risk of anxiety (p=0.003).

Chi-square and man Whitney -U Test were done to identify association between GH treatment and other confounding variables as gender and PWS genetic subtype, age of start of GH, and duration of GH treatment. On multivariate regression analysis age of start of GH treatment was an independent predictor for depression (OR=0.939,95% CI: 0,895-0.984, P=0.009). Duration of GH treatment was calculated based on age of start and stoppage of GH treatment. For those who still on GH treatment we used the last follow up visit date to calculate the duration of GH treatment. Duration of GH treatment not significantly associated with decrease rate of depression or increase in anxiety. Among participants with ages from 3-12 years, 60 % started GH treatment at mean age of 1.5±2 years. For those aged 12-18 years 50 % started GH treatment at age 6±4.4 years, and for participants >18 years 16% started GH treatment at age of 22 ± 14 years.

## Discussion

The aim of the study was to investigate the effect of molecular classes, age groups, sex, and GH treatment on behavioral disorders in PWS and the analyses presented here include results from one of the largest studies to date of the correlation between genotype and ASD in patients with PWS. The prevalence of ASD in PWS patients in our study was 19.5%, in concordance with previous studies at 25-41% (Veltman et al.,2015; Bennett et al.,2015). Dykens et al.,2017 reported that 12.3% of patients with PWS met the criteria for ASD diagnosis based on comprehensive assessments including the Autism Diagnostic Observation Schedule-2 (ADOS-2) and expert clinical evaluations. In our study participants with deletions are non-significantly associated with a higher risk for compulsive counting, compulsive ordering, visual hallucinations, and depressed mood. These findings are supported by the literature, which reports that patients with deletions have higher rates of developing compulsive behavior (Manzardo et al., 2018; Krefft et al.,2014). Patients with mUPD have been found to have less skin picking and maladaptive behavior (Dykenset al., 1999; Ymons et al., 1999). Sinnema et al.,2011 reported that patients with mUPD had more behavioral problems. In contrast to other studies, we found that Patients with deletions had higher frequencies of anxiety and skin picking than those with UPD. We categorized our patients into the three age groups, we found that Visual hallucinations, depression, and skin picking were significantly higher in the older age group. In agreement with our results Sinnema et al.,2011; Clarke et al., 1996; Whitman & Accardo, 1987, found that behavioral problems were more prevalent in adolescence and young adulthood. Young adulthood can be a time of psychosocial adaptations leading to increase of behavioral problems, compared to childhood and adolescence. Transition from school to work and feeling different in abilities and dependency compared with siblings, may lead to more maladaptive behavior in the adulthood (Boer et al.,2002). Systematic review by Bennet et al., 2015 showed that there is lack of studies on ASD in young patients with PWS as these symptoms become more evident with age. Most of the studies showed an average age of diagnosis above 8 years which is consistent with our results.

Based on reports that growth hormone may improve cognition and behavior in individuals with PWS, we speculated that growth hormone use would contribute to a decreased risk of psychiatric behaviors. GH treatment was significantly associated with lower rates of depression. GH treatment was associated with significant decrease in skin picking in PWS patients with deletion. Anxiety had a significant association with growth hormone use. However, early initiation of GH treatment was associated with a lower risk of depression without increasing risk of anxiety. Whitman et al., 2002 reported that GH treatment was associated with major reduction in depressive symptoms in patients older than 11 years old. No apparent behavioral deterioration associated with GH treatment. Eiholzer et al, 1998 found that GH treated PWS children were more attentive, independent, and less anxious. Absence of behavioral deterioration could be a considered a positive outcome for GH treatment.

Our study has some limitations, the exact age of onset of psychiatric symptoms was not consistently recorded. Detailed information on the age of onset of psychiatric symptoms as well as duration and frequency of episodes should also be included in the analyses. This information would be important in determining when symptoms started in relation to growth hormone use. Behavioral and psychiatric diagnosis was based on subjective evaluation by parent reports and data obtained through medical records rather than from a formal validated psychiatric testing. Increasing the sample size to include more adolescent and adult individuals would also be beneficial in order to give the study greater statistical power over a wider age range of individuals with PWS.

## Data Availability

The data presented in this study are not publicly available but are available from the corresponding author on reasonable request.

## Notes

### Competing Interest Statement

The authors have declared no competing interest.

### Funding Statement

This research was funded by National institute of Health, as well as the Prader Willi Syndrome Association USA.

### Author Declarations

This study was approved by the human subjects committee at each participating institution (e.g., University of California Irvine (UCI) Institutional Review Board (IRB) protocol number 2007-5605)

